# Comparing methods collecting mucosal secretions and detecting SARS-CoV-2 spike IgA in three laboratories across three countries

**DOI:** 10.1101/2025.05.06.25327072

**Authors:** Oscar Bladh, Katherina Aguilera, Salma Sheikh-Mohamed, Jessica Nardulli, Disha Bhavsar, Dylan Fitzgerald, Gagandeep Singh, Lesley Ward, Giulio Kleiner, Gary Chao, Nina Greilert Norin, Tamás Pongrácz, Charles Gleason, Matilda Berkell, Mikael Åberg, Florian Krammer, Jennifer L. Gommerman, Charlotte Thålin, Viviana Simon

## Abstract

**Background:** Mucosal IgA is key in preventing severe acute respiratory syndrome coronavirus 2 (SARS-CoV-2) infections. Several mucosal vaccines are in development, and consistent methodologies assessing mucosal IgA are crucial for evaluation across clinical trials.

**Methods:** We compared SARS-CoV-2 ancestral spike-specific IgA and secretory IgA (SIgA) in nasal secretions and saliva from 20 adults enrolled at Danderyd Hospital, Stockholm, Sweden, and 23 adults enrolled at the Icahn School of Medicine at Mount Sinai, New York, USA. Nasal secretions were collected by Nasosorption® and nasal swabs, and saliva by passive drooling, Salivette®, and saliva swabs. Antibody levels were measured in all samples using an electrochemiluminescence assay (ECL) and two enzyme-linked immunosorbent assays (ELISAs).

**Findings:** Spike-specific IgA and SIgA levels measured by ECL correlated well with those measured by ELISA across nasal and saliva samples (range 0.42-0.94, p<0.01), except for saliva collected by saliva swabs yielding lower IgA concentrations and weaker correlations (range -0.21-0.27). Spike-specific IgA levels also correlated well across collection methods (range 0.7-0.9, p<0.0001), with a weaker correlation between saliva collected by passive drooling and saliva swab (r=0.55, p<0.001). Although antibody levels correlated well between nasal secretions and saliva collected by passive drooling or Salivette® (range 0.64-0.86, p<0.01), they were >3-fold higher in nasal secretions than in saliva (p<0.01).

**Interpretation:** This multi-center study demonstrates an overall good comparability between spike-specific IgA and SIgA across assays and collection methods, except for saliva swabs. Our findings suggest that nasal secretions may be preferable due to higher spike-specific IgA levels compared to in saliva.

## Introduction

Mucosal IgA antibodies in the airways are essential for preventing severe acute respiratory syndrome coronavirus (SARS-CoV-2) infections [1–3], and their generation relies on mucosal antigen encounter [4–6]. Given that the current SARS-CoV-2 vaccines are administrated by intra-muscular injection, they mostly elicit IgG antibodies. These antibodies are suitable for containing viral replication by neutralization as well as through engagement of both the complement system and innate immune cells. These vaccines, however, fail to elicit sufficient mucosal IgA responses to protect against infections and viral spread [7]. Several mucosal SARS-CoV-2 vaccines are currently in development, aiming for robust immunity in the airways [8 9]

Saliva and nasal secretions can be collected by a variety of methods, and there is currently no standardized approach for quantifying respiratory mucosal antibody levels. Nasal secretions can be collected by commercially available nasosorption devices [5 10 11] or by simply swabbing the anterior nostrils [12 13]. Saliva can be collected by passive drooling [14–16], as well as with collection devices such as cotton swabs or absorbent cotton rolls [17 18]. Enzyme linked immunosorbent assays (ELISA) are commonly used for detecting and quantifying antibody levels in blood, saliva and nasal secretions [19–21]. Alternatives such as assays based on an electrochemiluminescence (ECL) readout might offer enhanced sensitivity and specificity but are limited by their need for expensive platform equipment.

There are no correlations established between mucosal antibody levels quantified by these two widely used platforms to date. With numerous mucosal vaccines under evaluation in clinical trials [8 9], standardized methodologies for assessment of mucosal IgA responses are crucial for enabling comparisons across clinical studies and real-world data. Aiming to provide insights towards standardized methodologies for assessment of mucosal antibody responses, this study compared SARS-CoV-2 spike-specific IgA, secretory IgA (SIgA), and IgG levels in nasal secretions and saliva in 43 participants from two geographical sites, procured using five different collection methods and quantified by ECL, and two in-house ELISAs across three different laboratories.

## Methods

### Study population

A total of 43 participants were enrolled, 20 healthcare workers from the COMMUNITY cohort (Stockholm, Sweden) and 23 adults as part of the Clinical Sample Collection Protocol for Patients with Viral Infections (New York, USA), (**Table 1**, **Figure 1**). All participants provided written consent and permissions for biospecimen banking and sharing. Sample collection began in March 2024 and concluded in May 2024. Nasal secretions were collected by nasal swab and Nasosorption®, and saliva was collected by Salivette®, saliva swab and passive drool. All samples were collected at the same study visit.

**Figure 1.**
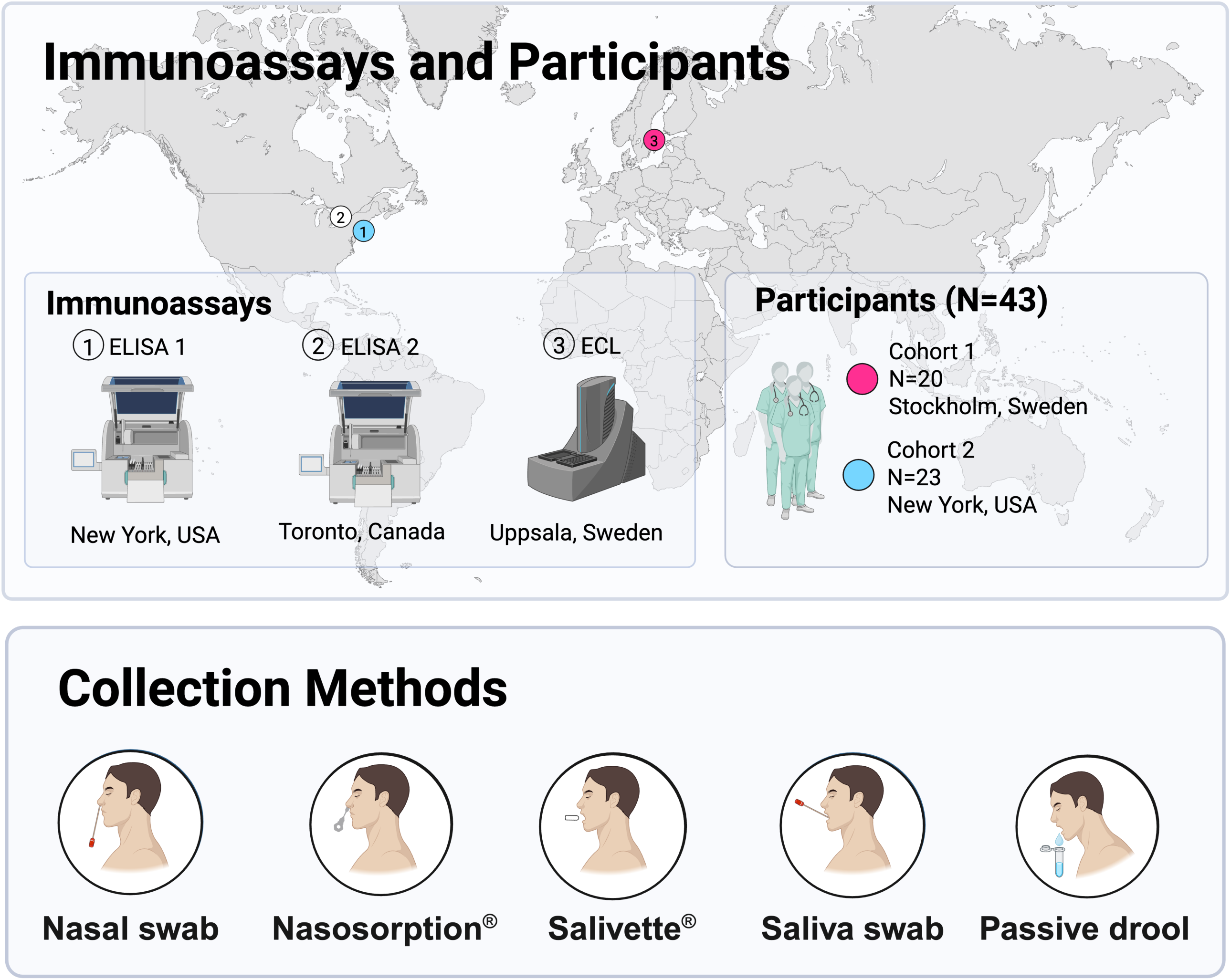
Study setup. Samples were collected from 43 adults in Sweden (Cohort 1, n = 20) and the USA (Cohort 2, n = 23). Five sample types were collected: nasal swab, Nasosorption®, Salivette®, saliva swab, and passive drool. Samples were analyzed using three different immunoassays targeting spike-specific IgA, SIgA, and IgG: two in-house ELISA methods (ELISA 1 and ELISA 2) and one commercial ECL platform. ELISA = Enzyme-linked immunosorbent assay; ECL = Electrochemiluminescence assay; SIgA = Secretory IgA.

**Table 1.**

| Participant Metadata Summary |  |  |  |  |  |  |  |
| --- | --- | --- | --- | --- | --- | --- | --- |
| Demographics |  | All (N=43) |  | Stockholm Cohort 1 (N=20) |  | New York Cohort 2 (N=23) |  |
| Age | Mean | 43 |  | 49 |  | 38 |  |
|  | Range | 21-69 |  | 21-64 |  | 24-69 |  |
|  |  | N | % | N | % | N | % |
| Sex at Birth | Male | 15 | 35% | 4 | 20% | 11 | 48% |
|  | Female | 28 | 65% | 16 | 80% | 12 | 52% |
| SARS-CoV-2 Vaccination and Infection History |  |  |  |  |  |  |  |
|  |  | N | % | N | % | N | % |
| Primary Vaccine Type | Pfizer | 22 | 51% | 13 | 65% | 9 | 39% |
|  | Moderna | 14 | 33% | 0 | 0% | 14 | 61% |
|  | Astra Zeneca | 4 | 9% | 4 | 20% | 0 | 0% |
|  | No vaccine | 3 | 7% | 3 | 15% | 0 | 0% |
| First SARS-CoV-2 Immune Exposure | Infection | 14 | 33% | 11 | 55% | 3 | 13% |
|  | Vaccination | 26 | 60% | 6 | 30% | 20 | 87% |
|  | N/A | 3 | 7% | 3 | 15% | 0 | 0% |
| Number of SARS-CoV-2 Infections | 0 | 5 | 11% | 0 | 0% | 5 | 22% |
|  | 1 | 21 | 49% | 7 | 35% | 14 | 61% |
|  | 2 | 10 | 23% | 7 | 35% | 3 | 13% |
|  | 3 | 2 | 5% | 1 | 5% | 1 | 4% |
|  | 4 | 2 | 5% | 2 | 10% | 0 | 0% |
|  | N/A | 3 | 7% | 3 | 15% | 0 | 0% |
| Number of SARS-CoV-2 Vaccine Doses | 0 | 3 | 7% | 3 | 15% | 0 | 0% |
|  | 3 | 19 | 44% | 11 | 55% | 8 | 35% |
|  | 4 | 8 | 19% | 4 | 20% | 4 | 17% |
|  | 5 | 10 | 23% | 1 | 5% | 9 | 39% |
|  | 6 | 3 | 7% | 1 | 5% | 2 | 9% |
| Number of total SARS-CoV-2 Immune Events (Vaccine, Infection) | 1 | 3 | 7% | 3 | 15% | 0 | 0% |
|  | 3 | 2 | 5% | 0 | 0% | 2 | 9% |
|  | 4 | 7 | 16% | 2 | 10% | 5 | 22% |
|  | 5 | 12 | 28% | 7 | 35% | 5 | 22% |
|  | 6 | 9 | 21% | 0 | 0% | 9 | 39% |
|  | 7 | 4 | 9% | 3 | 15% | 1 | 4% |
|  | 8 | 3 | 7% | 2 | 10% | 1 | 4% |
|  | N/A | 3 | 7% | 3 | 15% | 0 | 0% |

### The COMMUNITY cohort

The COMMUNITY cohort study (NCT06784739) is an ongoing observational study exploring immune responses to SARS-CoV-2 infection and vaccination [28–30]. Briefly, 2,149 healthcare workers (HCW) have been monitored with follow-ups conducted every four months, including the collection of serum, saliva, and nasal secretions. Information on vaccination type and timing is obtained from the Swedish vaccination register (VAL Vaccinera). SARS-CoV-2 infection is determined based on the fulfillment of any of the following criteria: a positive PCR test registered in the national communicable diseases register, seroconversion to the SARS-CoV-2 spike antigen during any follow-up before vaccination, seroconversion to the SARS-CoV-2 nucleocapsid antigen at any follow-up, or documentation of a positive rapid diagnostic test reported during any follow-up in the study. Written informed consent was obtained from all study participants and the study protocol was approved by the Swedish Ethical Review Authority (dnr 2020-01653).

### Clinical Sample Collection Protocol for Patients with Viral Infections

This study is an ongoing observational longitudinal study (IRB-17-00791/STUDY-16-01215) that collects samples from adults exposed to or with confirmed documented viral infection [31–34]. Upon enrollment and sample collection, complete viral infection and vaccination histories are recorded as provided by participants and confirmed where available by the electronic medical record and vaccination records. Participants with a wide range of immune histories and conditions have been observed, and both blood and mucosal samples have been banked. All participants gave written informed consent prior to collection of data and biospecimen. The study was approved by approved by the Program for the Protection of Human Subjects at the Icahn School of Medicine at Mount Sinai.

### Sample collection methods

The collection and processing of nasal and saliva samples followed standardized protocols. To ensure similar standard collection procedures between the two collection sites, a movie tutorial was implemented and used by the study staff performing the samplings in the study [35].

### Nasal secretions

*Nasal swab;* Nasopharyngeal flocked swab (Copan #CP503CS01) was gently inserted 2 cm centimeters into one nostril and rotated 5–6 times against the nasal mucosa. The swab was then placed into a 15 mL centrifuge tube containing 0.5 mL phosphate buffered saline (PBS), and the lid was securely sealed. The procedure was repeated for the other nostril, using a separate swab and tube. In the lab, tubes were vortexed for 10 seconds, and swabs were gently pressed against the inner wall of the tube to extract liquid before being discarded. Samples from the same participant were pooled.

*Nasosorption^®^;* Nasosorption*^®^* FX-i 7 mm nasal strip (Hunt Developments UK Ltd) was gently inserted into one nostril until resistance was met, with participants instructed to press their index finger against the side of the nostril to keep the strip in place for 60 seconds. The strip was then carefully removed and returned to its holder. The procedure was repeated for the other nostril, and the strips were kept on ice until further processing. Each nasal strip was then removed from its holder and placed into an Eppendorf tube containing 500 μL of an assay buffer containing PBS, 1% bovine serum albumin (BSA), and 0.05% Tween 20. Tubes were vortexed for 30 seconds, and the strips were gently pressed against the wall of the tube using a filter before being placed back into the tube. The tubes were centrifuged at 1,600xg for 20 minutes at 4°C, after which the strips and filters were discarded. Samples from both nostrils were pooled.

### Saliva

#### Salivette^®^

Participants were instructed to keep the Salivette^®^ absorbent roll (Sarstedt #51.1534) against the cheek for two minutes, occasionally alternating sides and gently chewing to stimulate saliva production. The roll was then returned to the Salivette^®^ tube. Samples were processed by centrifuging the tube at 1,000xg for 5 minutes at 21°C, after which the cotton roll and filter were removed and discarded.

#### Saliva swab

Two nasopharyngeal flocked swabs (Copan #CP503CS01) were immersed in saliva briefly without brushing the tongue or oral mucosa. Each swab was placed into a 15 mL centrifuge tube containing 0.5 mL PBS, tubes were then vortexed for 10 seconds, and the swabs were gently pressed against the inner wall of the tube to extract saliva before being discarded. Samples from the same participant were pooled.

#### Passive drool

*P*articipants provided saliva samples by passively drooling into a clean collection cup for five minutes.

The final steps were consistent across all collection methods; 400 μL of the extracted fluid was pipetted into two 0.5 mL cryovials, 50 μL into a 96-well plate (Sarstedt #83.3926), and the remaining volume into a 0.5 mL cryovial. All samples were then sealed and stored at -80°C, with plates labeled by sample type.

### Immuno Assays

#### ECL

All sample types were analyzed at a 1:100 dilution using the spike-specific IgA and IgG V-PLEX Respiratory Panel 4 according to the manufacturer’s instructions (Meso Scale Diagnostics (MSD), Maryland, USA). Spike-specific SIgA was measured using the same panels as the spike-specific IgA with the following modifications: A set of samples with anticipated high spike-specific SIgA levels was initially analyzed, and the three samples with the highest IgA signals were pooled to serve as calibrators for the standard curve. A four-fold serial dilution was prepared with two replicates at seven levels, plus a buffer-only zero-calibrator blank. After calibrator and sample incubation (sample dilution 1:50) followed by washing steps, the supplied detection antibody was replaced with a conjugated monoclonal mouse anti-human IgA antibody (HP6141, Calbiochem, Sigma-Aldrich) at a final concentration of 1 μg/mL and the final steps were completed according to the manufacturer’s protocol. The conjugation of the detection antibody was performed by Meso Scale at no cost. Total IgA and Total IgG concentrations were quantified at 1:5000 and 1:500 dilutions, respectively, using the Human/NHP IgA kit and the Human/NHP IgG kit (MSD), according to the manufacturer’s instructions.

An electrochemiluminescence reader was used to analyze all plates (MESO SECTOR S600 instrument, MSD). By correcting for dilution, the final antibody concentrations in undiluted samples were calculated and delivered as AU/mL for spike-specific IgA and spike-specific SIgA and pg/mL for total IgA. To account for possible differences in sampling, spike-specific IgA and spike-specific SIgA levels were normalized to total IgA levels in the same sample. The ratio was then multiplied by 10^7^ for graphical purposes.

#### ELISA 1

ELISA to measure IgA, IgG and SIgA against SARS-CoV-2 spike protein were performed as described in detail previously [16 19] For a detailed protocol please consult [36]. Briefly, Immulon-4 HBX 96-well microtiter plates (Thermo Fisher Scientific) were coated with SARS-CoV-2 spike protein diluted to 2 μg/mL in PBS overnight at 4°C. Well contents were discarded and plates were blocked with 200 μL/well of 5% non-fat milk in PBS containing 0.1% Tween 20 (PBS-T) for one hour at room temperature (RT). After blocking, 50 μL of samples (starting at 1:2 dilution followed by two-fold serial dilution in 2.5% non-fat milk in PBS-T) were added to the plates. An SIgA version of CR3022 monoclonal antibody [37] was used as a positive control. For IgA measurement, the incubation was carried out for two hours at RT. After 3 washes with 200 μL/well of PBS-T, 50 μL of goat anti-Human IgA (α-chain specific)-horeseradish peroxidase (HRP) conjugated (A0295, Sigma-Aldrich) diluted to 1:3000 was added to each well and incubated at RT for 1 hour. For IgG measurement, the reaction of samples with antigen was conducted at RT for 2 h. After washing with PBST three times, 50 μL of HRP-labeled goat anti-human IgG (H + L) antibody (Fisher Scientific, #PI31412) diluted to 0.32 μg/mL with 2.5% non-fat milk in PBST was added to each well, and incubated at RT for 1 h. For SIgA measurement, the plates were incubated with the samples overnight at 4°C. After washing with PBS-T, 50 μl of mouse anti-human secretory IgA antibody (HP6141, MilliporeSigma) diluted to 5 μg/mL was added to each well and incubated at RT for 2 hours. The plates were washed again with PBS-T and 50 μL of goat anti-mouse IgG Fc antibody-HRP conjugated (31439, Invitrogen) was added to each well and incubated at RT for 1 hour. Following incubation with HRP-labelled antibody, all set of plates, were washed with PBS-T and 100 μL of SIGMAFAST o-phenylenediamine dihydrochloride (OPD) substrate solution (Sigma-Aldrich) was added to each well. Ten minutes after incubation with OPD solution at RT, 50 μL of 3N hydrochloric acid (HCl) was added to each well to stop the reaction and the optical density (OD) at 490nm was measured using Synergy4 microplate reader (BioTek). Baseline for each plate was determined by averaging the OD value of blank wells and adding 3 times the standard deviation. Binding curves for the samples were generated and area under the curve (AUC) values were calculated using GraphPad Prism 10, taking the baseline values into account. Adjusted AUC values were obtained by dividing the AUC values by the total IgA concentration for the sample.

#### ELISA 2

ELISA were conducted for the detection of anti-spike IgA, IgG and secretory component as previously described [3]. Briefly, streptavidin-coated 96-well plates (ThermoFisher, 436014) were coated using 50 µL per well of biotinylated spike antigen (20ug/mL in PBS). Control wells were coated with PBS only. After overnight incubation at 4°C, the antigen-coated plates were blocked with a 5% BLOTTO (5% w/v skim milk powder in PBS) solution for 2 hours at 37°C. Frozen saliva samples were thawed and diluted in 2.5% BLOTTO at a range of 1:5-1:20. Next, diluted samples were pre-incubated on a separate streptavidin-coated plate for 30 minutes at 37°C to reduce anti-streptavidin activity before being transferred to the antigen-coated wells for a 2-hour incubation at 37°C. After washing 3x with PBS-Tween 20, 50 μL of HRP-conjugated goat anti-human-IgG and IgA antibodies (Southern Biotech, IgG: 2044-05, IgA: 2053-05) and sheep anti-human secretory component antibody (Vector labs, SHAHU/SC/PO) were added to the appropriate wells at dilutions of 1:1000, 1:2000 and 1:750 in 2.5% BLOTTO, respectively, and incubated for 1 h at 37°C. Plates were then washed 3x with PBS-Tween 20, followed by development using 50 μL per well of 3,3′,5,5′tetramethylbenzidine (TMB) Substrate Solution (ThermoFisher, 00-4021-56). Development was stopped by adding 50 µL/well of 1 N H_2_SO_4_, and OD was read at a wavelength of 450 nm (OD450) on a spectrophotometer (Thermo Multiskan FC). Adjusted OD values (value of sample well minus value of PBS-control well) were used to calculate the AUC for each sample.

### Statistics

Antibody levels are presented as medians with interquartile ranges and were compared using the Mann-Whitney U test or, when applicable, the Kruskal-Wallis test with Dunn’s post hoc test for multiple comparisons. Chi-Square tests were conducted to assess statistical differences between observed and expected preferences for each collection method category. Correlations were assessed using Spearman’s rank correlation. The coefficient of variation (%CV) was calculated as %CV = (standard deviation/mean) * 100. All statistical analyses were performed in GraphPad Prism version 9.2.0 (GraphPad Software, San Diego, California, USA) or R version 4.4.2.

When comparing non-normalized spike-specific Ig levels across different assays, samples with no detectable spike-specific IgA values but with a detectable corresponding total IgA level were assigned a value of 10^-5^. This adjustment ensures these data points are retained in the calculations, as the detection of total IgA indicates the presence of IgA in the sample, even if spike-specific IgA is undetectable. Similarly, adjustments were applied to normalized spike-specific Ig values when the specific Ig value was undetectable, but the corresponding total Ig value was measurable. In such cases, normalized spike-specific IgA and spike-specific secretory IgA values were assigned 10^-3^. Assigned values were chosen to be below the lowest detectable spike-specific IgA level (10^-5^), or the below the lowest detectable normalized spike-specific IgA and spike-specific SIgA values (10^-3^), in the dataset, reflecting the lack of direct measurement while preserving the informational value of these samples. These adjustments facilitate comparison across assays while maintaining the integrity of the dataset.

### Role of funders

None of the funding sources had any role in study design, data analysis, preparation of manuscript or decision to submit the manuscript for publication.

## Results

### Study design

Nasal secretions and saliva samples were collected from 20 adults enrolled at Danderyd Hospital, Stockholm, Sweden (Cohort 1), and 23 adults enrolled at the Icahn School of Medicine at Mount Sinai, New York, USA (Cohort 2), **(Table 1).** Nasal secretions were collected by Nasosorption®, and FLOQ swab (hereafter nasal swab). Saliva was collected by passive drooling, Salivette®, and FLOQ swab (hereafter saliva swab). All samples were analyzed using two in-house ELISAs (hereafter referred to as ELISA 1 and ELISA 2), and on an ECL platform, for the presence of SARS-CoV-2 spike-specific IgA, SIgA and IgG (**Figure 1**). Correlations between antibody levels measured by ECL and the ELISAs, between collection devices within the same mucosal compartment and between mucosal compartments were assessed.

### Correlations between spike-specific IgA levels measured by ECL and two ELISAs in nasal secretions and saliva

We first aimed to address intra-individual correlations between ancestral spike-specific IgA and SIgA measured in nasal secretions and saliva using ECL and the two ELISAs.

Spike-specific IgA levels measured by ECL correlated with those measured by both ELISAs in samples collected from both the nasal and oral compartments (r range from 0.61-0.94, p<0.001), except for saliva samples collected by saliva swab where the correlation was weaker between ECL and ELISA 2 (r=0.27, p=0.09). Similar correlations were observed between spike-specific SIgA measured by ECL and ELISA 1 (r range from 0.63-0.91, p<0.001). However, spike-specific SIgA measured by ECL and ELISA 2 generally displayed a weaker correlation (r range from 0.42-0.67, p<0.01), where no correlation was observed in saliva samples collected by saliva swab (r= -0.21, p=0.17) (**Figure 2A-B, Figure S1**). Correlations between spike-specific IgG levels across all assays are shown in **Figure S2**, and correlations of spike-specific IgA and SIgA levels between ELISA1 and ELISA 2 are demonstrated in **Figure S3**.

**Figure 2.**
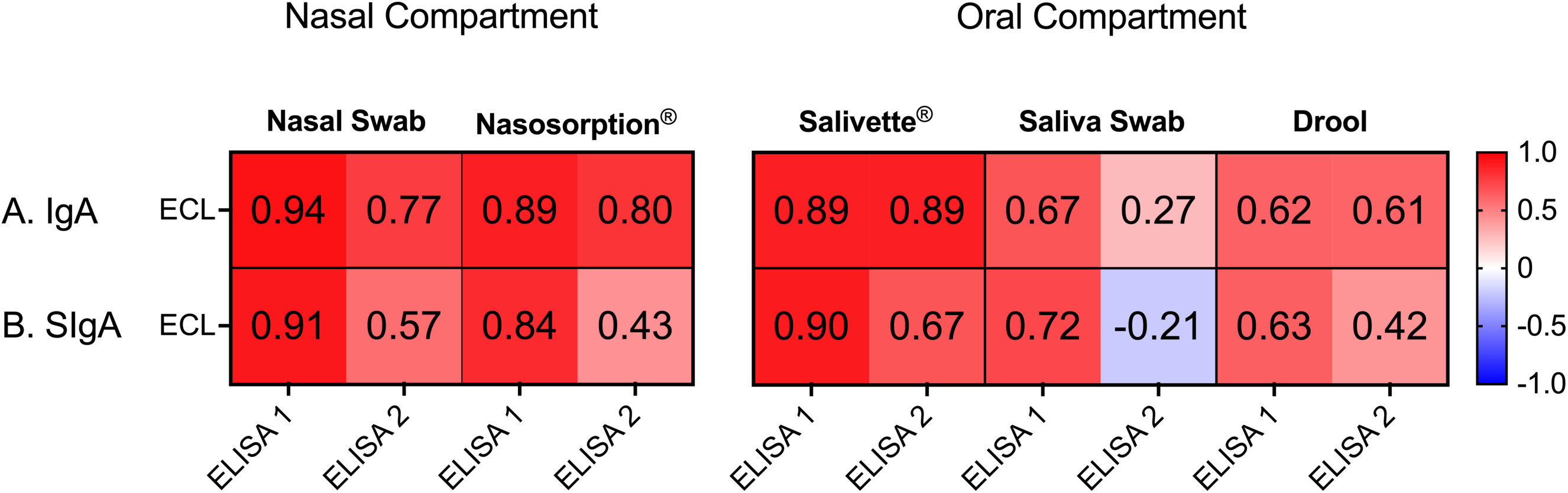
Heatmap of correlation coefficients between spike-specific IgA (A) and spike-specific SIgA (B) across the nasal and oral compartment using three different assays. Correlations between assays: ECL (commercial electrochemiluminescence assay, SciLifeLab), ELISA 1 (in-house ELISA, Krammer laboratory), and ELISA 2 (in-house ELISA, Gommerman laboratory). IgA: spike-specific immunoglobulin A; SIgA: spike-specific secretory IgA. Correlation strength is color-coded, with red indicating positive correlations and blue indicating negative correlations. Spearman’s rank correlation was used to determine correlation coefficients between assays. ECL = Commercial electrochemiluminescence assay, SciLifeLab, ELISA 1: In-house ELISA, Krammer laboratory, ELISA 2: In-house ELISA, Gommerman laboratory. IgA: Spike-specific IgA, SIgA: Spike-specific secretory IgA.

Collectively, spike-specific IgA and spike-specific SIgA levels measured by ECL and the two ELISAs correlated well in both nasal secretions and saliva, except for saliva samples collected by saliva swabs.

### Impact of collection device on spike-specific IgA levels in nasal secretions and in saliva

Next, we assessed whether the collection device affected spike-specific IgA levels quantified by ECL in nasal secretions and in saliva by determining intra-individual correlations between spike-specific IgA levels collected by various sampling methods. Spike-specific IgA levels were normalized to total IgA levels in the same sample. We chose to compare results obtained by the commercially available ECL assay as it has the highest sensitivity and is produced under ISO certification (Meso Scale Diagnostics (MSD), Maryland, USA). Accordingly, our technical replicates across all sample types indicated good precision, with the following percent coefficient of variation (%CV): nasal swab, 8.0; Nasosorption®, 6.7; Salivette®, 9.3; saliva swab, 12.9; and passive drool, 6.2. Moreover, correlations between spike-specific IgA and specific SIgA, analysed with ECL, across all collection methods, were very strong (R>0.9) (**Figure S4**), which suggest that IgA can be used as a proxy for SIgA in mucosal secretions.

Correlations were high between spike-specific IgA levels measured by the various collection devices, both in nasal secretions and in saliva (r:0.7-0.9, p<0.0001), except for a weaker correlation between spike-specific IgA in saliva collected by passive drooling and saliva swab (r=0.55, p<0.001). However, there was a notable difference in correlations between saliva collection devices when stratifying the results between the two cohorts (r: 0.77–0.92, p<0.0001 in Cohort 1, vs. r: 0.46–0.52, p<0.05 in Cohort 2, with the exception of r=0.07, p=0.75, for the correlation between passive drool and saliva swab) (**Figure 3A-C**). In contrast, correlations in the nasal compartment were higher and comparable between the two cohorts (r: 0.80–0.96, p<0.0001, **Figure 3D**). As expected, there were significant differences in total IgA levels between collection devices, with the lowest levels in saliva swabs (**Figure 3E**). However, after normalization against total IgA levels in each sample, spike-specific IgA levels were similar in nasal secretions collected by nasal swabs and Nasosorption® device (median 71.6 vs 80.6 AU/mL) and in saliva collected by Salivette®, saliva swab and passive drool (medians 18.7 vs 22.3 vs 25.3 AU/mL, **Figure 3F**). Correlations for the same analysis on ELISA 1 and ELISA 2 are shown in **Figure S5**.

**Figure 3.**
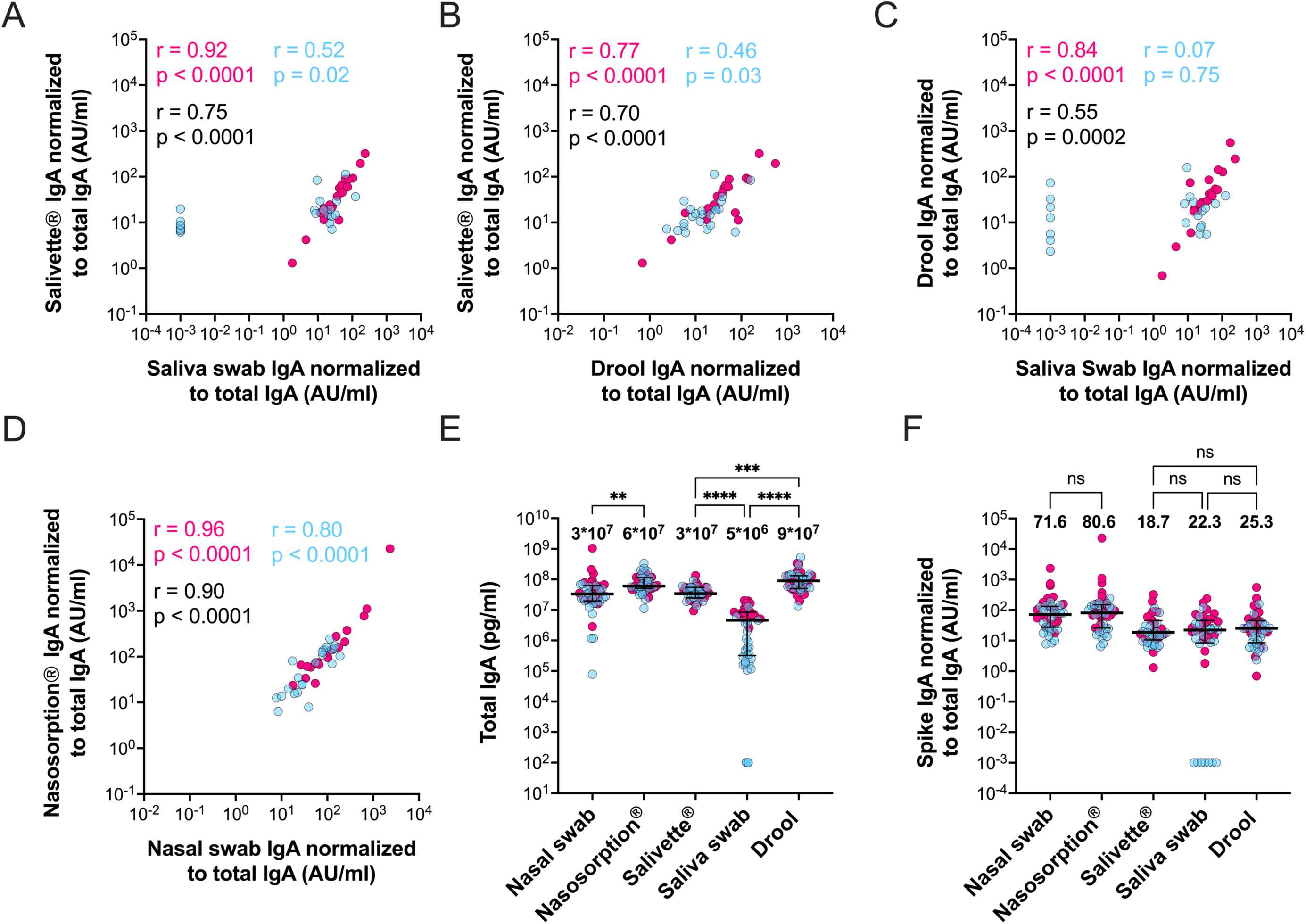
Correlations between normalized spike-specific IgA levels within the oral and nasal compartments, and total and normalized spike-specific IgA levels stratified by collection method. Correlations between normalized spike-specific IgA levels across oral (Salivette®, saliva swab, drool) and nasal (nasal swab, Nasosorption®) compartments (A-D). Correlation coefficients (r-values) and p-values are shown on each graph. Total IgA levels (E) and spike-specific IgA levels normalized to total IgA levels (F), stratified by collection method. Median antibody levels are displayed above each sampling method. Sample sites are color-coded as follows: pink (Cohort 1), blue (Cohort 2). Horizontal black lines indicate median values with interquartile ranges. Statistical significance was determined using the Kruskal– Wallis test with Dunn’s multiple comparisons correction. Significance levels between sampling methods are indicated by horizontal brackets. Spearman’s rank correlation was used to calculate correlation coefficients. (ns = not significant; ** p < 0.01; *** p < 0.001; **** p < 0.0001). AU/mL = Arbitrary Units per milliliter; pg/mL = picograms per milliliter.

These comparisons demonstrate that antibodies can be reliably measured in nasal secretions collected by either a Nasosorption® device or by nasal swabs, and in saliva using either Salivette® or passive drooling. However, saliva collected with saliva swabs poses challenges due to low total IgA concentrations. Nevertheless, with a sensitive assay, these samples are comparable in terms of spike-specific IgA after normalization to total IgA.

### Participant-reported preference for different sample types

To assess potential discomfort associated with each collection method, a subset of participants from cohort 1 (n=14) were asked to rate their experience using a Numeric Rating Scale (NRS) ranging from 0 (’no discomfort’) to 10 (’maximum discomfort’). The collection methods were ranked by median discomfort levels as follows: Salivette® (4.5/10), Nasosorption (2.5/10), nasal swab (2/10), passive drool (2/10), and saliva swab (0.5/10). Salivette® was associated with significantly greater discomfort compared to saliva swab (p<0.05) (**Figure S6**). Overall, saliva swab emerged as the most preferred collection method (selected by 58% of participants, p<0.01) and was particularly favoured among saliva collection methods (86% chose saliva swab, p<0.01). Among nasal secretion collection methods, nasal swab was preferred over Nasosorption® by 71% of participants (p = 0.09) (**Figure S7**). While most collection methods were associated with only slight discomfort, Salivette® was rated significantly more uncomfortable than the other methods.

### Comparison of spike-specific Ig levels in nasal secretions and saliva

Finally, we compared spike-specific IgA and spike-specific SIgA levels between the nasal and oral mucosal compartments. All samples were analyzed on the ECL platform, and spike-specific IgA and spike-specific SIgA levels were normalized against total IgA levels in the same sample to account for differences in sample concentration.

There were strong correlations between IgA levels in both nasal secretions and saliva collected by either device (**Figure 4A-B**). Notably, levels of spike-specific IgA and spike-specific SIgA were significantly higher in nasal secretions as compared to saliva, **Figure 4C-D**. Collectively, this demonstrates that while spike-specific IgA and spike-specific SIgA levels are higher in nasal secretions than in saliva, they correlate well, except for in saliva collected by saliva swab. Scatterplots of spike-specific IgA, SIgA and IgG correlations between oral and nasal compartments analyzed with ECL are depicted in **Figure S8** and analyzed with ELISA 1 and ELISA 2 are depicted in **Figure S9** and **Figure S10** respectively.

**Figure 4.**
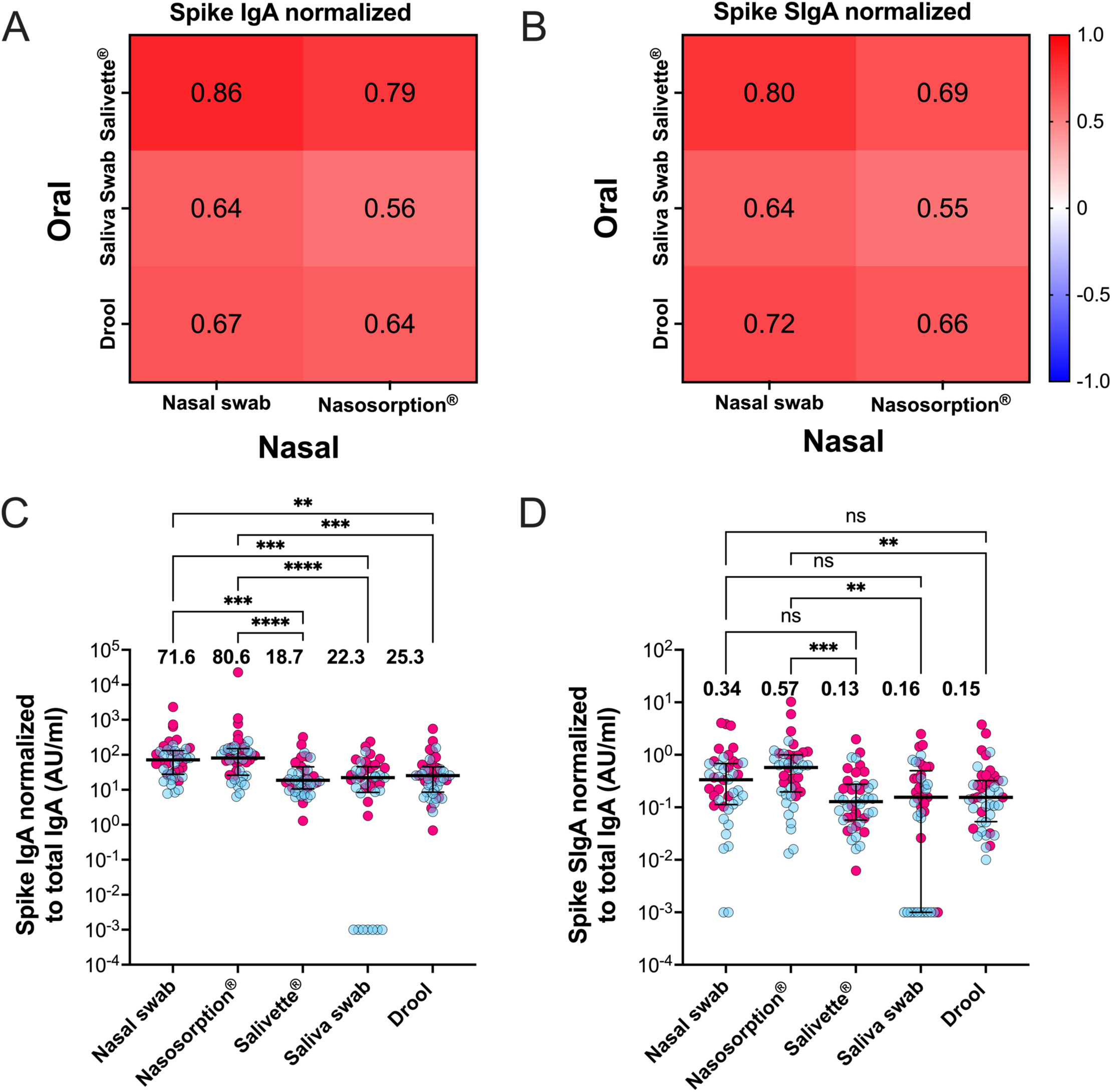
Comparison of oral and nasal antibody levels. Correlations between oral (Salivette®, saliva swab, drool) and nasal (nasal swab, Nasosorption®) compartments for spike-specific IgA (A) and spike-specific SIgA (B), both normalized to total IgA levels. Correlation coefficients (r-values) are shown in heatmaps, with red indicating positive correlations and blue indicating negative correlations. Scatter plots showing spike-specific IgA (C) and spike-specific SIgA (D), normalized to total IgA levels, across different sampling methods. Horizontal black lines indicate median values with interquartile ranges. Median antibody levels are displayed above each scatter plot. Sample sites are color-coded as follows: pink (Cohort 1) and blue (Cohort 2). Statistical significance was determined using the Kruskal– Wallis test with Dunn’s multiple comparisons correction. Spearman’s rank correlation was used to calculate correlation coefficients. Significance levels between sampling methods are indicated by horizontal brackets. (ns = not significant; ** p < 0.01; *** p < 0.001; **** p < 0.0001). SIgA: spike-specific secretory IgA.

## Discussion

This study compared mucosal spike-specific antibody levels measured by different collection methods and quantified by three different immunoassays. We found an overall good comparability between spike-specific IgA and SIgA across assays and collection methods. However, although spike-specific IgA and SIgA levels correlated well between nasal and oral compartments, they were considerably higher in nasal secretions compared to saliva, which is in line with our previous findings [12].

Our comparisons revealed strong correlations between spike-specific IgA and SIgA levels measured by the ECL and ELISA platforms, with a few notable exceptions. Whereas SIgA levels correlated well between ELISA 1 and ECL, correlation between the levels of SIgA quantified by these assays and ELISA 2 were weaker. Both ECL and ELISA 1 employed antibodies detecting secretory IgA, while ELISA 2 incorporated an antibody targeting the secretory component. The secretory component is a proteolytic cleavage product of the polymeric IgG receptor (pIgR) which remains attached to both IgA and IgM following pIgR-mediated transcytosis across the mucosal epithelium [22 23]. The weaker correlation between SIgA measured with ELISA 2 and the other two assays may therefore mirror a lower specificity for SIgA by ELISA 2 due to this particular secondary detection antibody. Given that SIgA and IgA correlate well (Figure S4), measuring IgA only may be sufficient if one were to choose a single readout. The other exception was that saliva swabs appeared to be less robust compared to other oral collection methods, as demonstrated by the large variation in salivary swab IgA concentrations between sample sites in comparison to other oral collection methods. The lower antibody concentrations due to the higher dilutions of saliva swabs as compared to passive drooling or Salivettes may furthermore pose challenges for less sensitive assays.

Strong SARS-CoV-2 spike-specific antibody level correlations were demonstrated between collection devices within the same mucosal compartment after normalization to total IgA levels in the same samples to account for factors such as sample dilution, stress and circadian rhythm that may affect the levels of secreted IgA [24]. Exceptions here included correlations between antibody levels in samples collected by saliva swab and the other oral collection methods when quantified by ELISA. However, these correlations remained strong when quantified by ECL, suggesting that the low antibody concentrations in samples collected by saliva swabs may still be reliably quantified if antibody levels are analyzed with a sensitive assay and normalized to total IgA level in the same sample. Saliva swabs and nasal swabs furthermore emerged as the most preferred collection methods in the participant-reported preference survey, which could be important to consider in larger studies, where the ease of the sampling process is essential to ensure continued participation [12].

Finally, we corroborate our prior findings by showing that normalized spike-specific IgA levels were consistently higher in the nasal compartment compared to the oral compartment [12]. IgA levels in external secretions are influenced by many factors that need consideration when interpreting these results [25]. In saliva, periodontal inflammation and salivary flow rate may influence the proportion of secreted IgA [26], and recent breakthrough infection can also increase total IgA levels [27]. Theoretically, differences in the density of resident spike-specific IgA-secreting plasma cells between the two compartments could also explain the observed variation in spike-specific IgA secretion, possibly due to prior viral exposure or physiological factors. However, to our knowledge, there is currently no established explanation for why the proportion of antigen-specific IgA is nearly twice as high in nasal secretions compared to saliva.

Our study has several limitations, including a relatively small sample size (43 adults provided 215 biospecimens and 14 NRS surveys). Moreover, we were not able to provide full evaluations of each assay due to limited biospecimen volumes. Most participants had experienced, at least, one SARS-CoV-2 infection prior to study participation, resulting in generally high spike-specific IgA levels. We were not able to evaluate how collection methods and assays perform when analyte levels are lower, but most adults today will have had a SARS-CoV-2 infection in the past five years.

Developing standardized protocols for sample collection, processing, and analysis is crucial to facilitate robust cross-study comparisons. As mucosal vaccines are being developed, consistent and reliable measurements of mucosal antibody responses are essential for evaluating mucosal immunogenicity and advancing our understanding of correlates of protection against upper respiratory infections. Our in-depth evaluation of five different methods for the collection of mucosal secretions in 43 adults enrolled at two different academic sites demonstrate that antibody levels correlate well between collection devices, assays as well as between the oral and nasal compartments. Moreover, our data suggest that nasal secretions may be the preferable compartment to sample from due to higher spike-specific IgA levels and consistent correlations across methods and geographic sample sites. Nasal swabs are particularly advantageous due to their ease of sampling, minimal discomfort, and simple preanalytical handling. Taken together, our findings have important implications for standardizing approaches evaluating mucosal immune responses across clinical studies.

## Data Availability

All data produced in the present study are available upon reasonable request to the authors

## Declaration of interest

The Icahn School of Medicine at Mount Sinai has filed patent applications relating to SARS-CoV-2 serological assays, NDV-based SARS-CoV-2 vaccines influenza virus vaccines and influenza virus therapeutics which list FK as co-inventor and FK has received royalty payments from some of these patents. VS is also listed on the SARS-CoV-2 serological assays patent. Mount Sinai has spun out a company, Kantaro, to market serological tests for SARS-CoV-2 and another company, Castlevax, to develop SARS-CoV-2 vaccines. FK is co-founder and scientific advisory board member of Castlevax. FK has consulted for Merck, GSK, Sanofi, Curevac, Seqirus and Pfizer and is currently consulting for 3rd Rock Ventures, Gritstone and Avimex. The Krammer laboratory is also collaborating with Dynavax on influenza vaccine development and with VIR on influenza virus therapeutics.

## Acknowledgements

We thank all the participants for their generous and continued support of our COVID-19 research programs. Special thanks go to the team of the Personalized Virology Initiative for processing and cryopreserving biospecimen from study participants.

This work was funded in part by the Centers of Excellence for Influenza Research and Surveillance (CEIRS, contract # HHSN272201400008C), the Centers of Excellence for Influenza Research and Response (CEIRR, contract # 75N93021C00014), by the Collaborative Influenza Vaccine Innovation Centers (CIVICs contract # 75N93019C00051), by philanthropic donations and by institutional funds including the Mount Sinai Center for Vaccine Research and Pandemic Preparedness, SciLifeLab & Wallenberg Data Driven Life Science Program, Knut and Alice Wallenberg Foundation, the National Bioinformatics Infrastructure Sweden (NBIS) at SciLifeLab, Region Stockholm and the Jonas and Christina af Jochnick Foundation.

## Data Sharing Statement

The data that support the findings of this study are available upon reasonable request from the corresponding authors. The data are not publicly available due to privacy of research participants.

## Supplementary appendix - Comparing methods collecting mucosal secretions and detecting SARS-CoV-2 spike IgA in three laboratories across three countries

**Figure S1.**
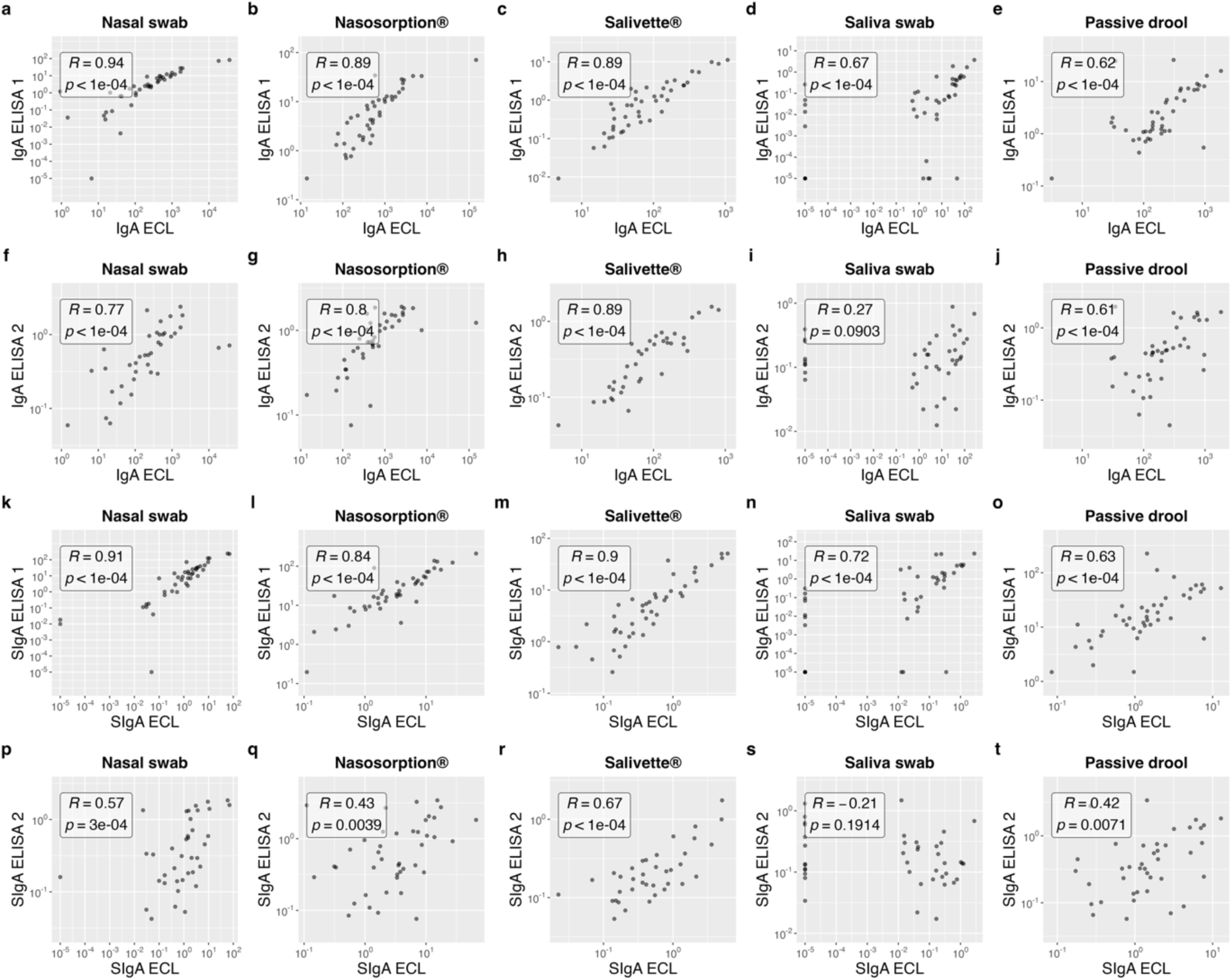
Correlations between spike-specific IgA and SIgA measured by ECL and ELISA in nasal secretions and saliva. Correlations between spike-specific IgA (a-j) and SIgA (k-t) levels measured by ECL and two in-house ELISA platforms (ELISA 1 and ELISA 2) across nasal secretions and saliva samples collected using nasal swab, Nasosorption®, Salivette®, saliva swab, and passive drool. Spearman’s rank correlation was used to calculate correlation coefficients (R), and p-values are indicated for each comparison. Axes represent spike-specific antibody levels in arbitrary units per milliliter (AU/mL). SIgA = spike-specific secretory IgA; ECL = electrochemiluminescence assay; ELISA = enzyme-linked immunosorbent assay.

**Figure S2.**
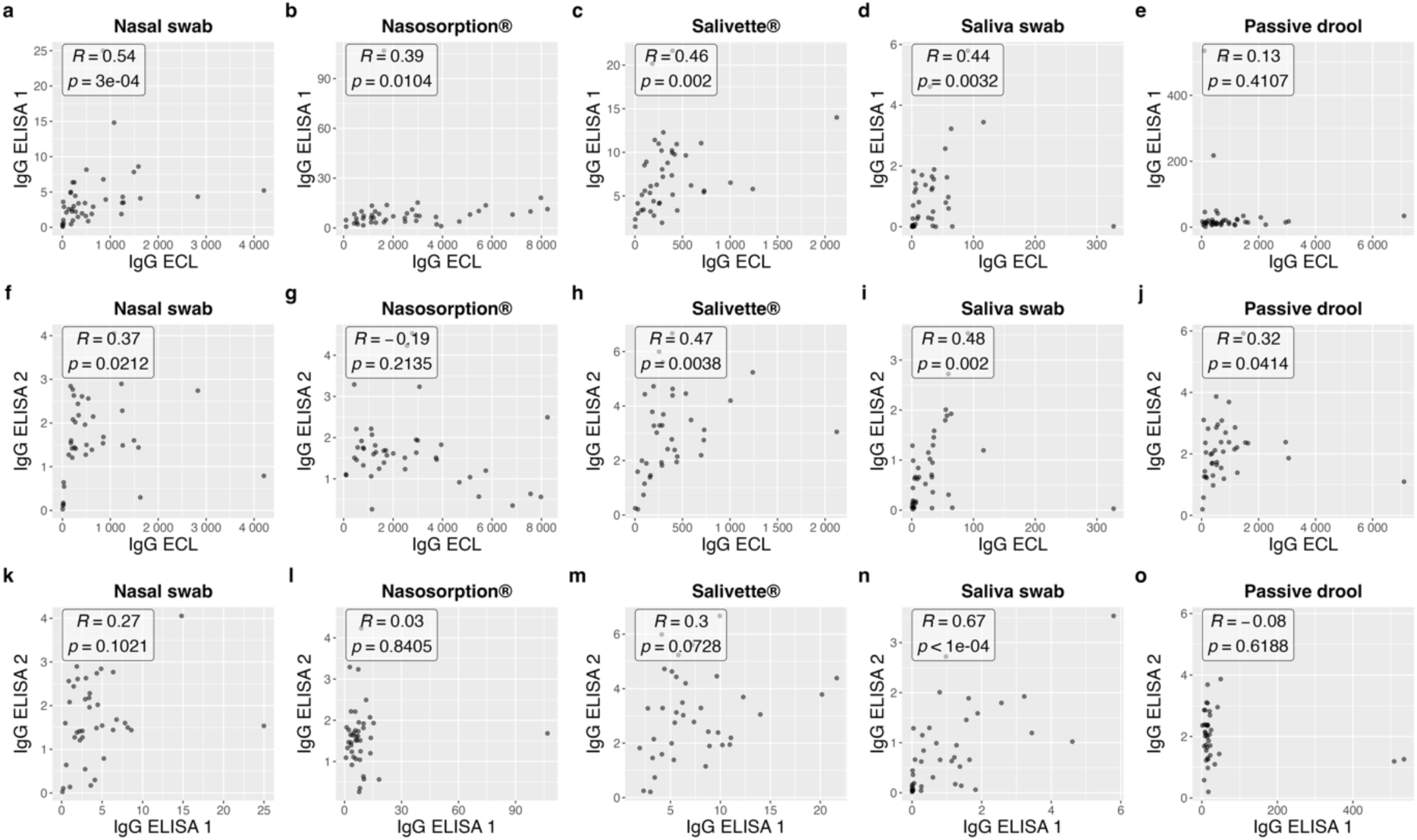
Correlations between spike-specific IgG measured by ECL and ELISA in nasal secretions and saliva. Correlations between spike-specific IgG levels measured by ECL and ELISA 1 (a-e), ECL and ELISA 2(f-j), and ELISA1 and ELISA 2 (k-o), across nasal and oral samples collected using nasal swab, Nasosorption®, Salivette®, saliva swab, and passive drool. Spearman’s rank correlation was used to calculate correlation coefficients (R), and p-values are indicated. Axes represent spike-specific antibody levels in AU/mL for ECL, and area under the curve (AUC) for ELISA 1 and ELISA 2.

**Figure S3.**
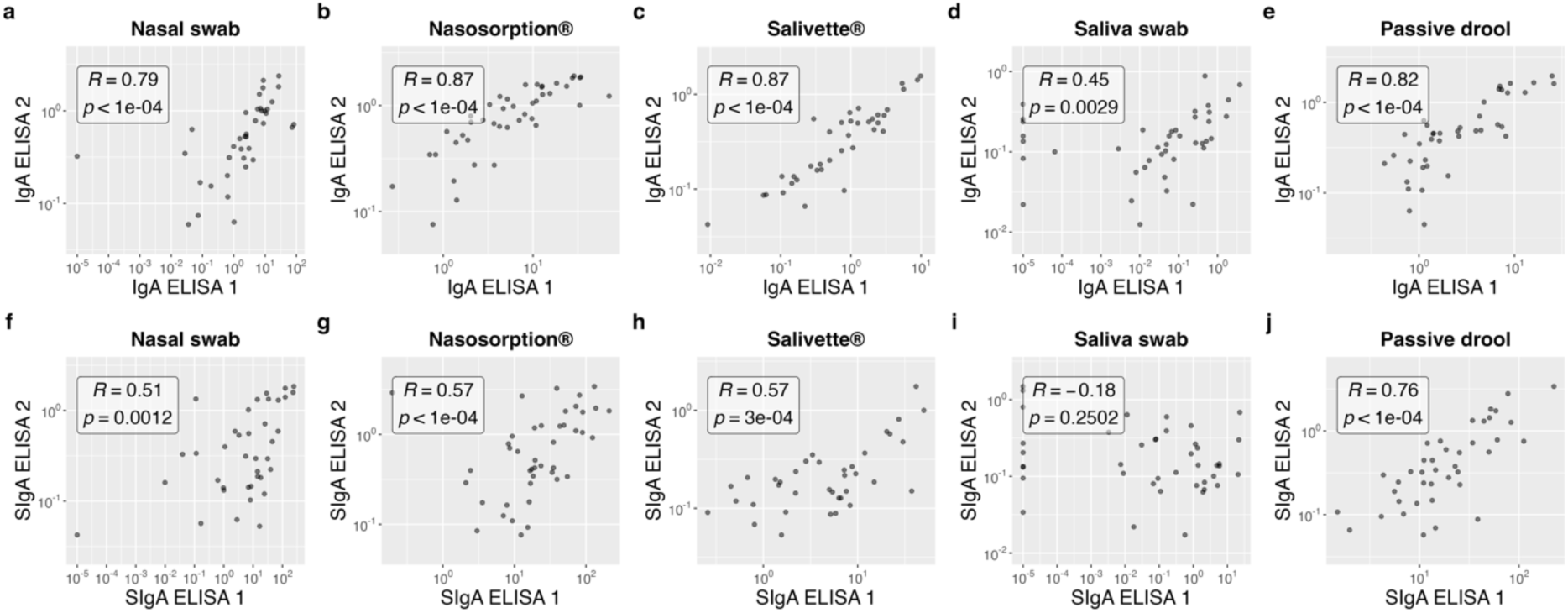
Correlations between spike-specific IgA and SIgA measured by ELISA 1 and ELISA 2 in nasal secretions and saliva. Correlations between spike-specific IgA (a-e) and SIgA (f-j) levels measured by ELISA 1 and ELISA 2 across nasal and oral samples collected using nasal swab, Nasosorption®, Salivette®, saliva swab, and passive drool. Spearman’s rank correlation was used to calculate correlation coefficients (R), and p-values are indicated. Axes represent spike-specific antibody levels in area under the curve (AUC). SIgA = spike-specific secretory IgA.

**Figure S4.**
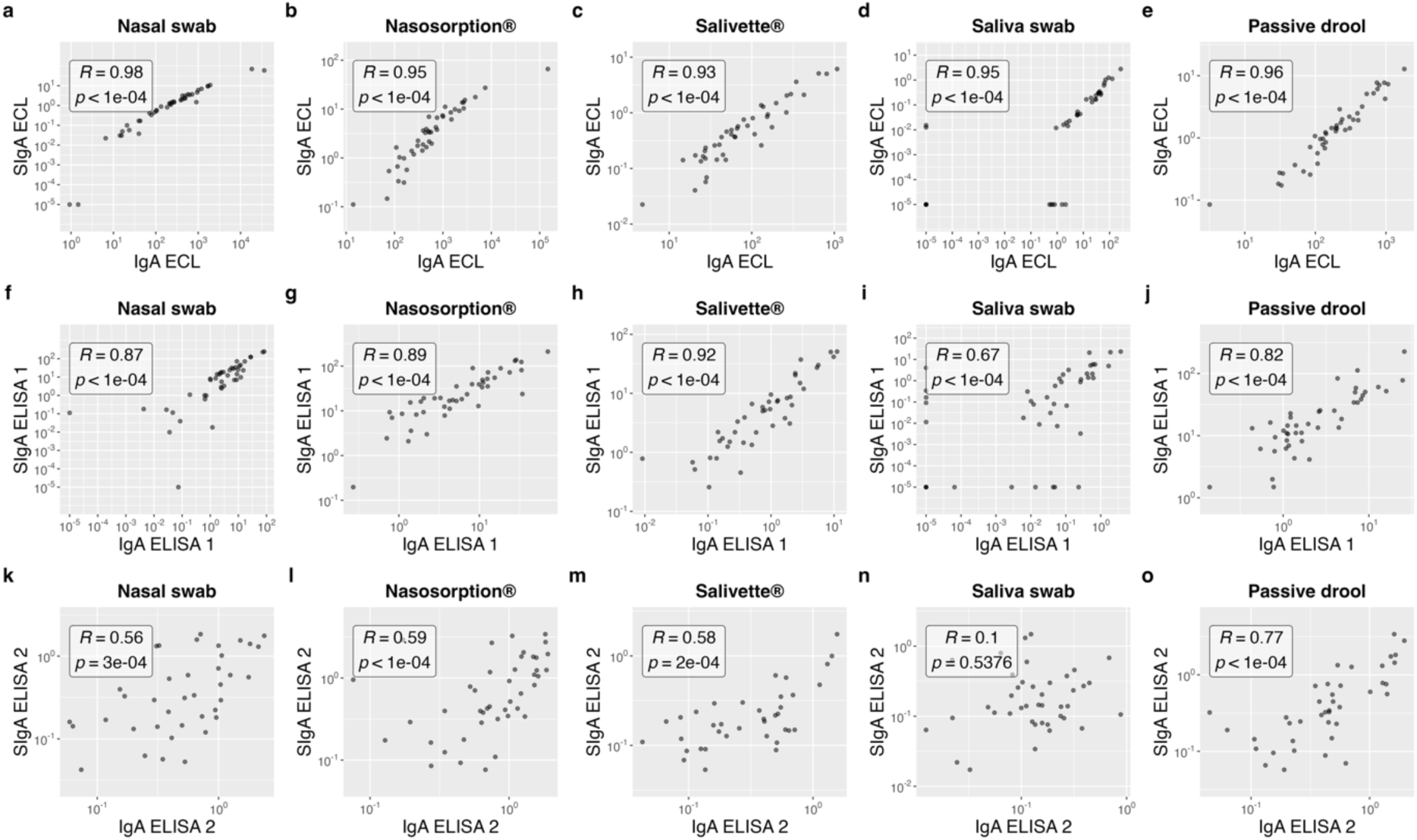
Correlations between spike-specific IgA and SIgA across ECL, ELISA 1, and ELISA 2 in nasal secretions and saliva. Correlations between spike-specific IgA and SIgA levels measured using ECL (a-e), ELISA 1 (f-j), and ELISA 2 (k-o) across nasal and oral samples collected using nasal swab, Nasosorption®, Salivette®, saliva swab, and passive drool. Spearman’s rank correlation was used to calculate correlation coefficients (R), and p-values are indicated. Axes represent spike-specific antibody levels in AU/mL for ECL and AUC for ELISA 1 and ELISA 2. SIgA = spike-specific secretory IgA.

**Figure S5.**
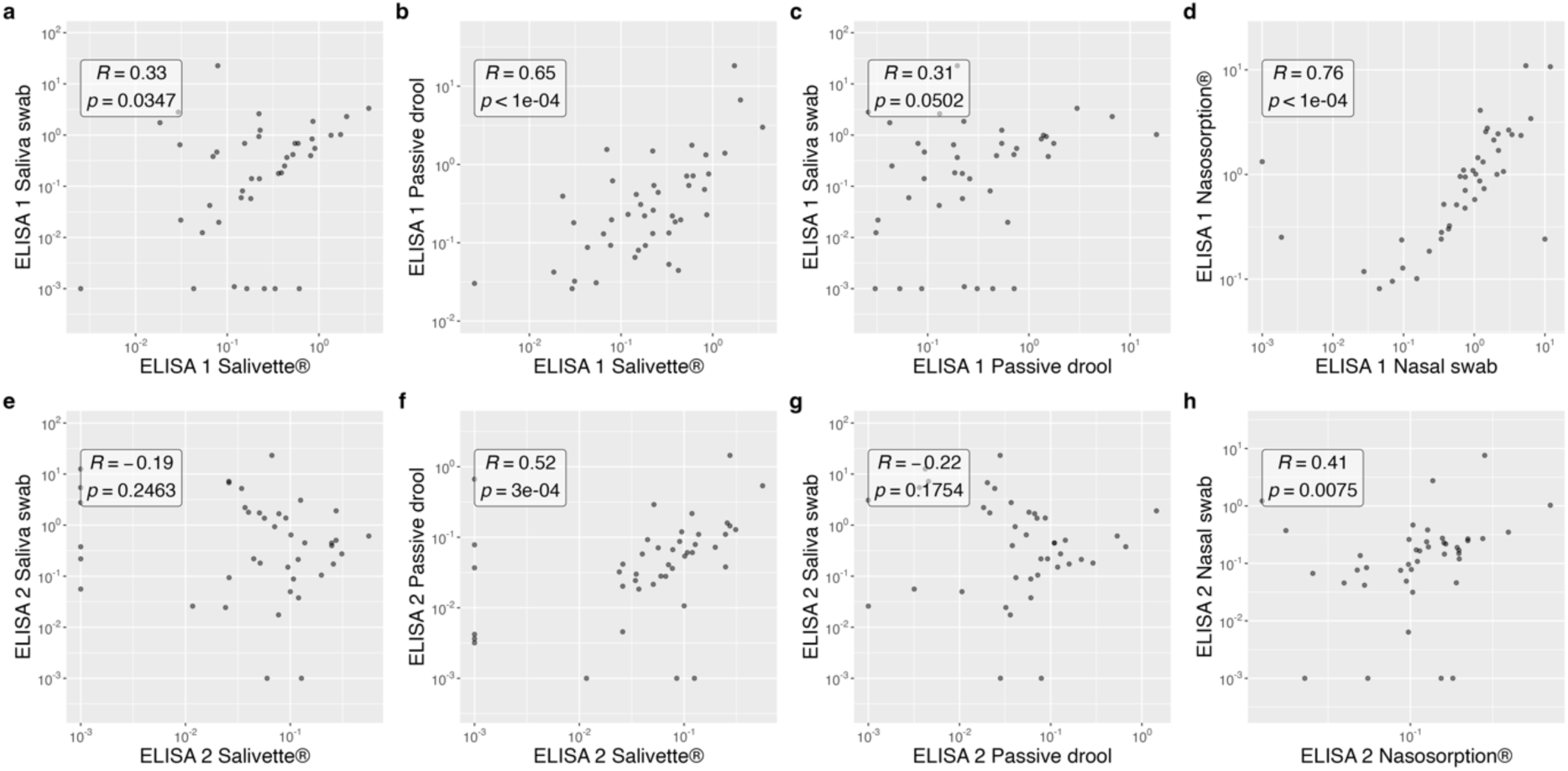
Correlations between normalized spike-specific IgA levels within the oral and nasal compartments analyzed by ELISA 1 and ELISA 2. Correlations between spike-specific IgA levels normalized to total IgA within the oral and nasal compartment, measured by ELISA 1(a-d) and ELISA 2 (e-h). Spearman’s rank correlation was used to calculate correlation coefficients (R), and p-values are indicated. Axes represent normalized spike-specific IgA values in AUC.

**Figure S6.**
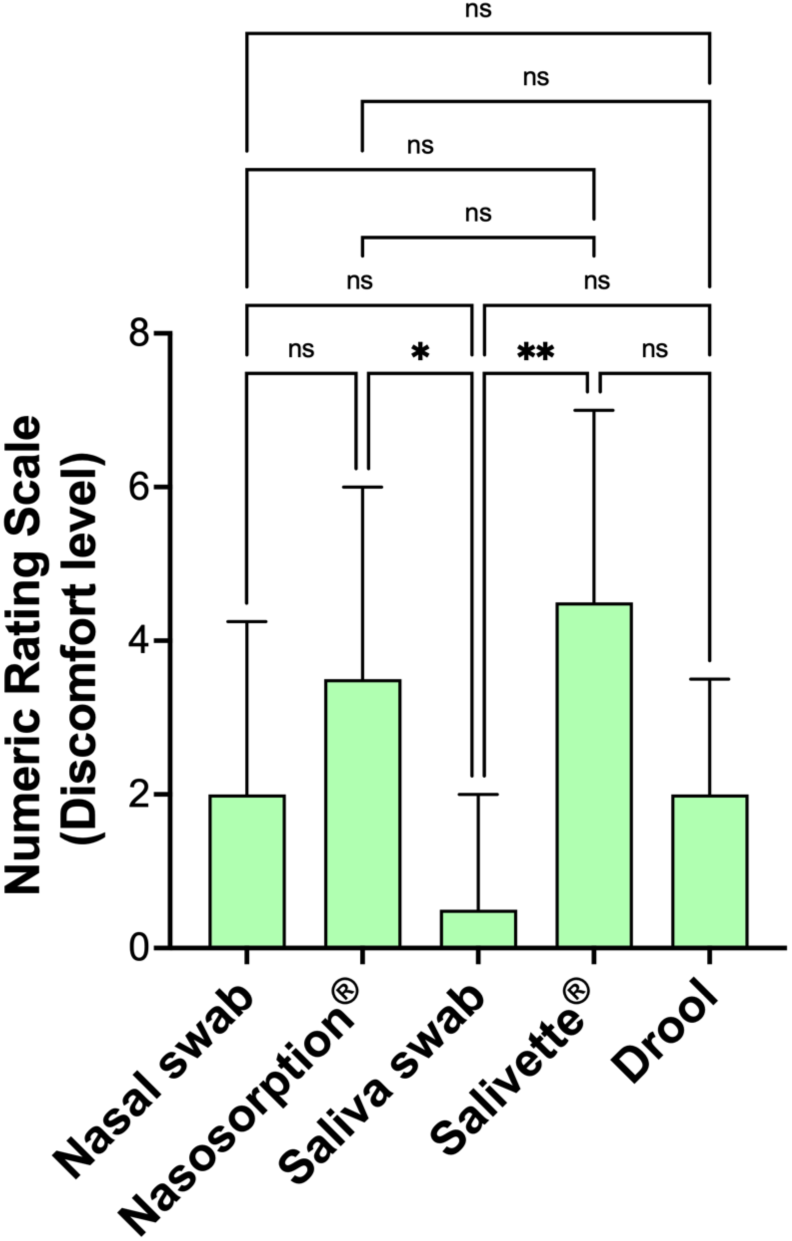
Participant-reported discomfort associated with different mucosal sampling methods. Bar chart showing median Numeric Rating Scale (NRS) scores from participants (n = 14) rating discomfort for each sampling method. Error bars indicate the 75th percentile. (ns = not significant; * p < 0.05; ** p < 0.01)

**Figure S7.**
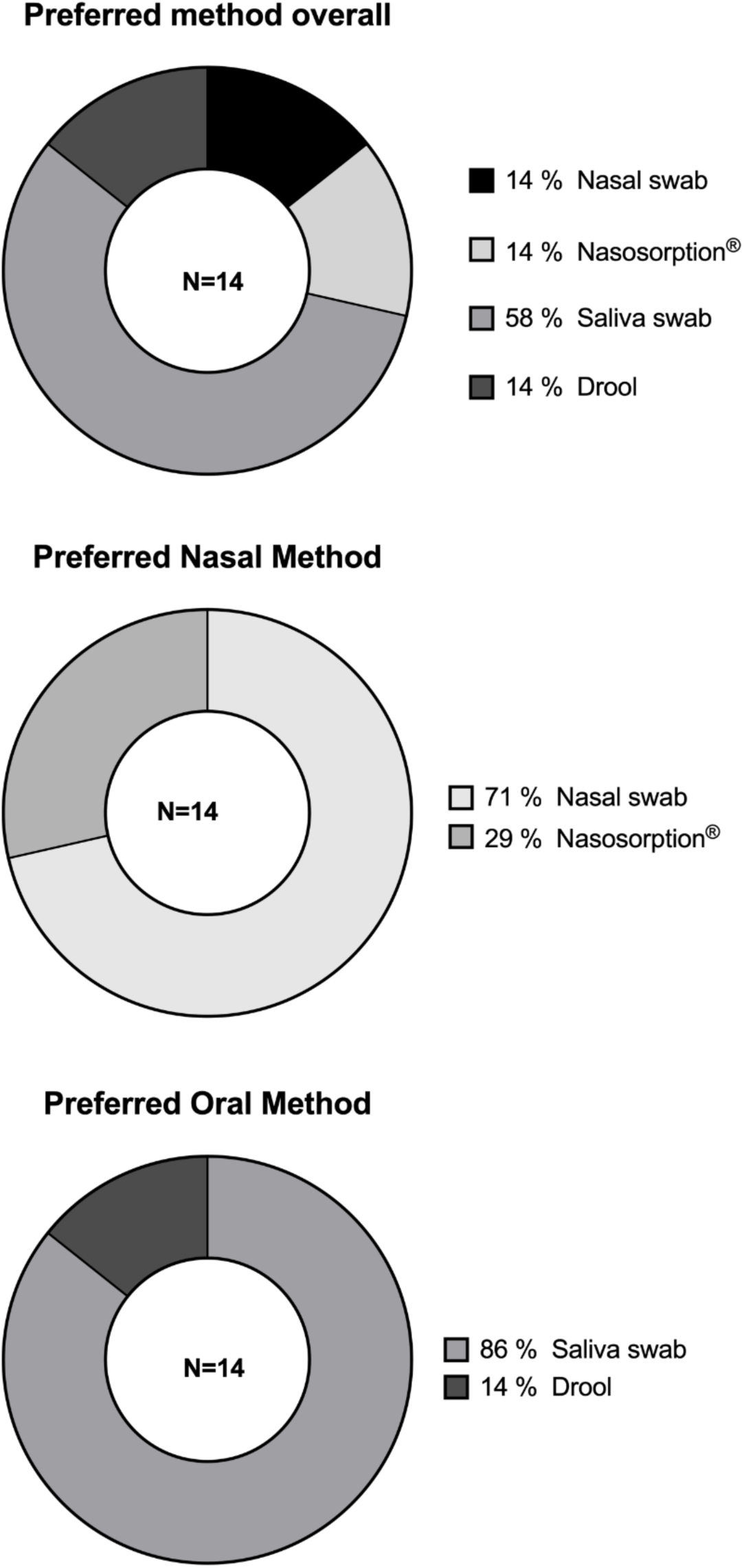
Participant preferences for mucosal sampling methods. Pie chart summarizing participants’ preferred collection methods for nasal and oral sampling.

**Figure S8.**
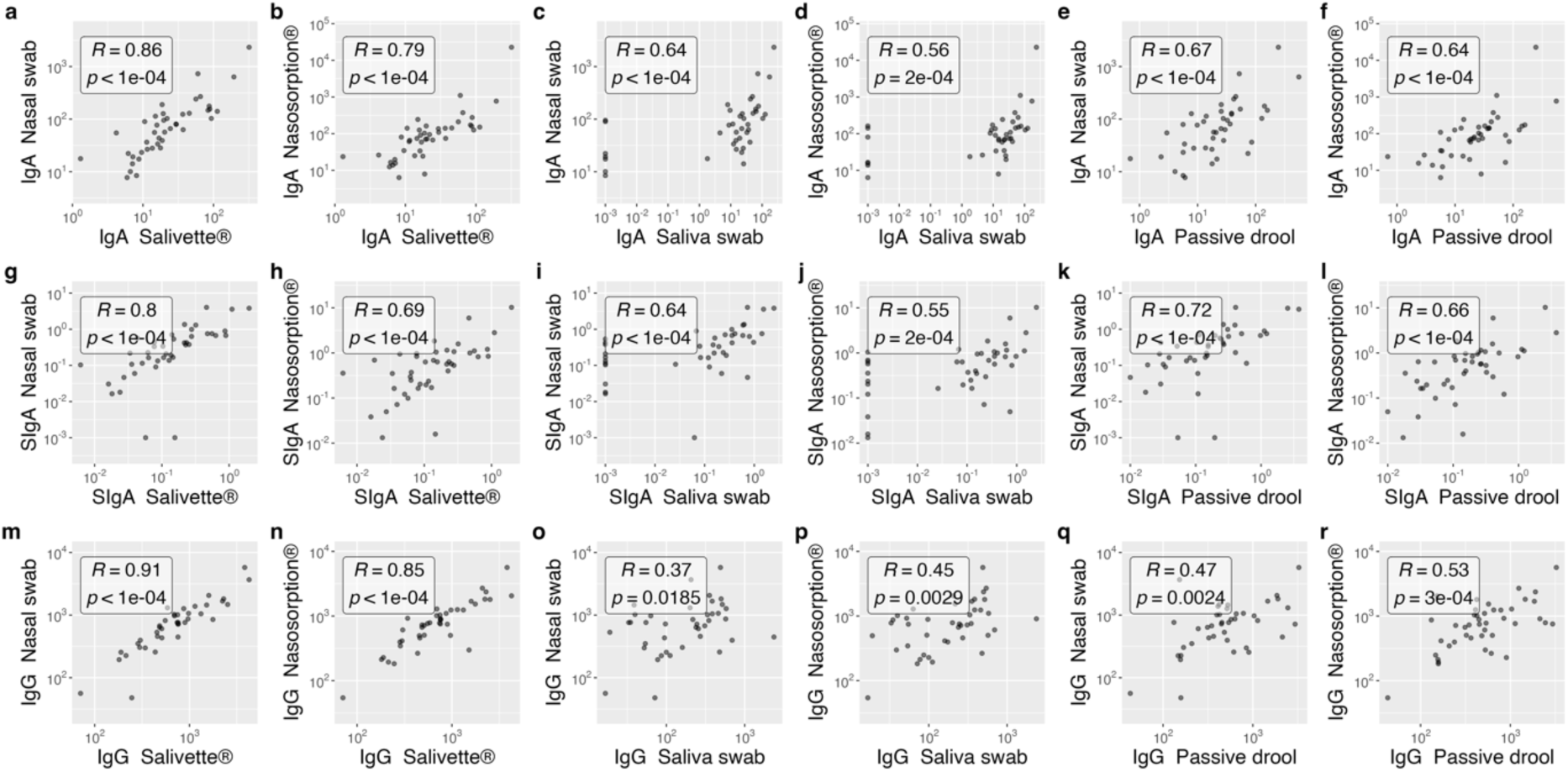
Correlations between nasal and oral sampling methods for spike-specific IgA, SIgA, and IgG measured using ECL. Correlations between nasal and oral collection methods for spike-specific IgA (a-f), SIgA (g-l), both normalized to total IgA, and IgG (m-r), normalized to total IgG, analyzed with ECL. Spearman’s rank correlation was used to calculate correlation coefficients (R), and p-values are indicated. Axes represent antibody levels in AU/mL. SIgA = spike-specific secretory IgA.

**Figure S9.**
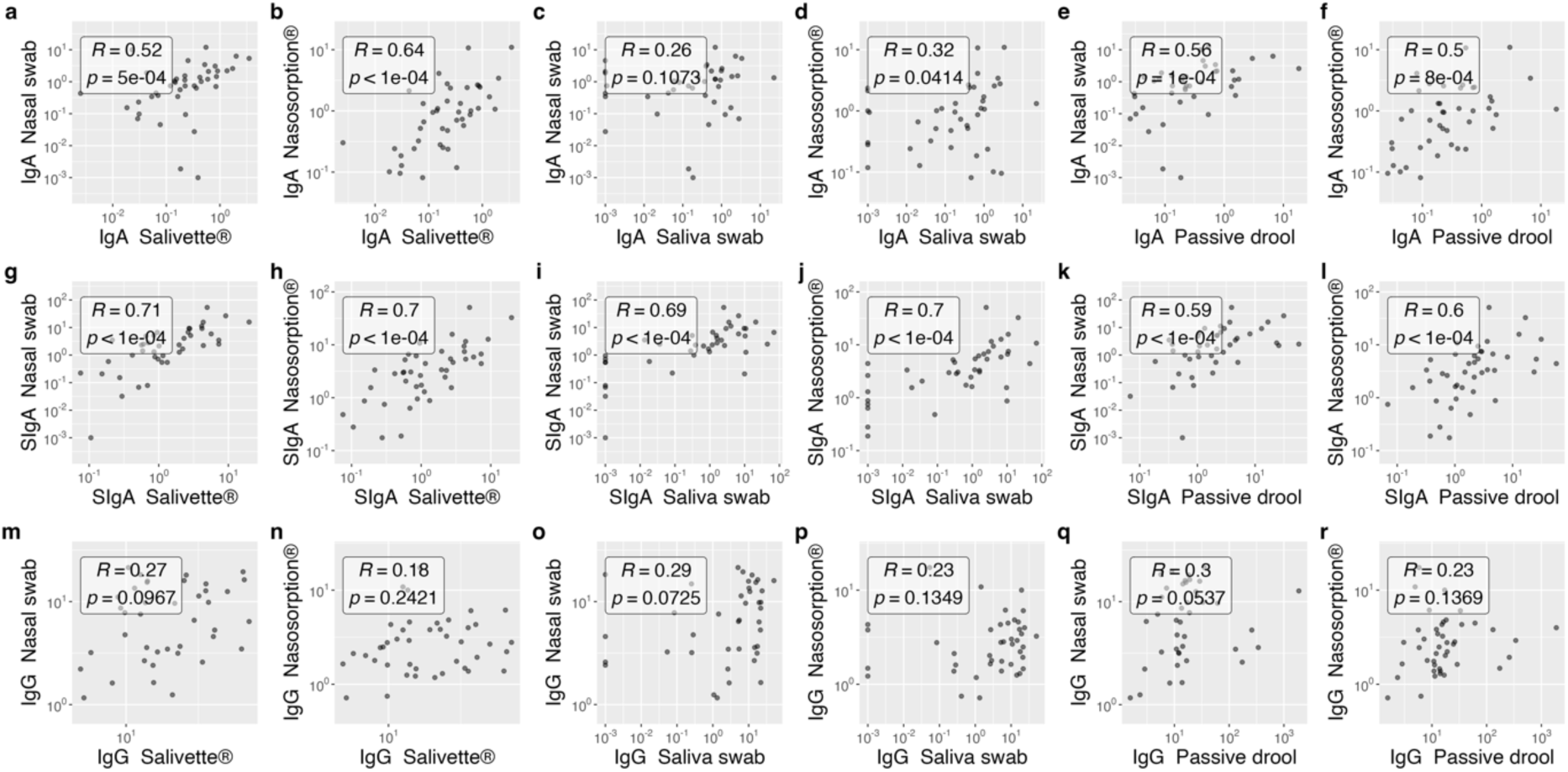
Correlations between nasal and oral sampling methods for spike-specific IgA, SIgA, and IgG measured using ELISA 1. Correlations between nasal and oral collection methods for spike-specific IgA (a-f), SIgA (g-l), both normalized to total IgA, and IgG (m-r), normalized to total IgG, analyzed with ELISA 1. Spearman’s rank correlation was used to calculate correlation coefficients (R), and p-values are indicated. Axes represent antibody levels in AUC. SIgA = spike-specific secretory IgA.

**Figure S10.**
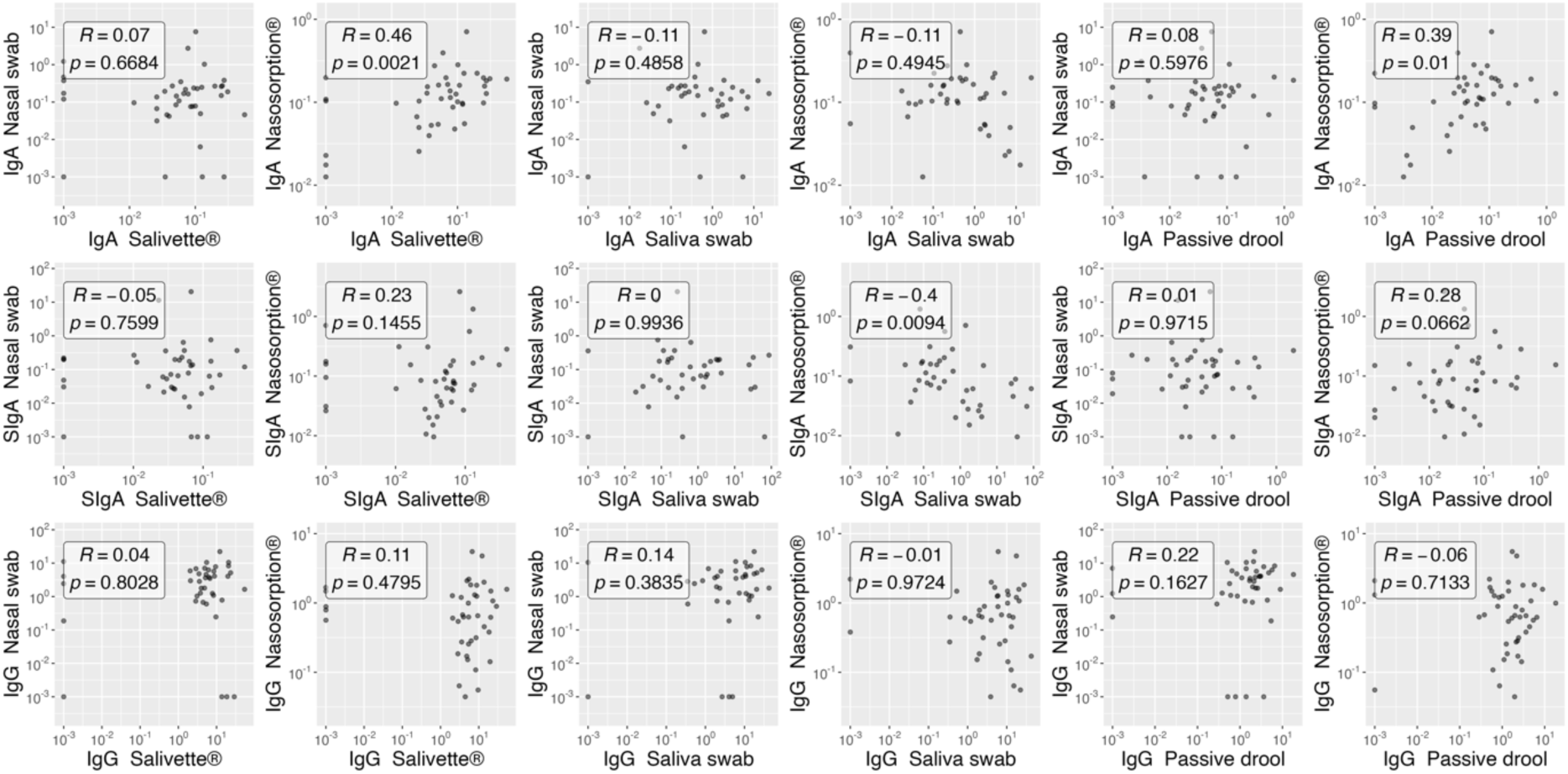
Correlations between nasal and oral sampling methods for spike-specific IgA, SIgA, and IgG measured using ELISA 2. Correlations between nasal and oral collection methods for spike-specific IgA (a-f), SIgA (g-l), both normalized to total IgA, and IgG (m-r), normalized to total IgG, analyzed with ELISA 2. Spearman’s rank correlation was used to calculate correlation coefficients (R), and p-values are indicated. Axes represent antibody levels in AUC. SIgA = spike-specific secretory IgA.

## Notes

### Author Declarations

Samples for this study were sourced from two onging, IRB approved observational studies: 1: an ongoing longitudinal study (IRB-17-00791/STUDY-16-01215) that collects samples from adults exposed to viral pathogens (Icahn School of Medicine at Mount Sinai). The Institutional Review Board of the Mount Sinai School of Medicine approved the Clinical Sample Collection Protocol for Patients with Viral Infections cohort (IRB-17-00791/STUDY-16-01215). -the COMMUNITY cohort study (NCT06784739), which is an ongoing study exploring immune responses to SARS-CoV-2 infection and vaccination. It has been approved by the Swedish Ethical Review Authority (dnr 2020-01653)

